# Pre-stroke physical activity matters for functional limitations: A longitudinal case-control study of 12,860 participants

**DOI:** 10.1101/2023.09.14.23295576

**Authors:** Zack van Allen, Dan Orsholits, Matthieu P. Boisgontier

**Affiliations:** School of Rehabilitation Sciences, Faculty of Health Sciences, University of Ottawa, Canada; Perley Health Centre of Excellence in Frailty-Informed Care, Ottawa, Canada; School of Human Kinetics, Faculty of Health Sciences, University of Ottawa, Canada

**Keywords:** Cohort Studies, Comorbidity, Disability, Exercise, Functional Status, Health Behavior, Longitudinal Studies, Prognosis, Prospective Studies, Stroke Survivors

## Abstract

**Objective:** In the chronic phase after a stroke, limitations in activities of daily living (ADLs) and instrumental ADL (IADLs) initially plateau before steadily increasing. The benefits of pre-stroke physical activity on these limitations remain unclear. To clarify this relationship, we examined the effect of physical activity on the long-term evolution of functional limitations in a cohort of stroke survivors and compared it to a cohort of matched stroke-free adults.

**Methods:** Longitudinal data from 2,143 stroke survivors and 10,717 stroke-free adults aged 50 years and older were drawn from a prospective cohort study based on the Survey of Health, Ageing and Retirement in Europe (2004-2022; 8 data collection waves). Physical activity was assessed in the pre-stroke wave. Functional limitations were assessed in the post-stroke waves. Each stroke survivor was matched with 5 stroke-free adults who had similar propensity scores computed on the basis of key covariates, including baseline age, sex, body mass index, limitations in ADL and IADL, chronic conditions and country of residence, before any of the participants from either cohort had experienced a stroke.

**Results:** Results showed an interaction between stroke status and physical activity on ADL limitations (b = -0.076; 95% CI = -0.142 to -0.011), with the effect of physical activity being stronger in stroke survivors (b = -0.345, 95% CI = -0.438 to -0.252) than in stroke-free adults (b = -0.269, 95% CI = -0.269 to -0.241).

**Conclusion:** The beneficial effect of pre-stroke physical activity on ADL limitations after stroke is stronger than its effect in matched stroke-free adults followed for a similar number of years.

**Impact:** Physical activity, an intervention within the physical therapist’s scope of practice, is effective in reducing the risk of functional dependence after stroke. Moreover, pre-stroke levels of physical activity can inform the prognosis of functional dependence in stroke survivors.

## INTRODUCTION

The prevalence of stroke exceeds 100 million cases worldwide^1^. On average, each of these cases is associated with a loss of 1.4 years of full health^1,2^. Over the past three decades, the number of years of full health lost to stroke has increased by an average of 1.2 million per year^1^. This burden on stroke survivors is reflected in their functional limitations. Specifically, one year after a stroke, survivors experience at least slight (59%)^3–17^, moderate (33%)^13–28^, or severe (23%)^11–13,15–20^ dependency in activities of daily living (ADLs), such as dressing, walking, bathing, eating, and toileting (Supplemental Tables S1 to S3). Regarding instrumental ADLs (IADLs), 40%^9,10,16,19,20^ of stroke survivors are moderately active and 41%^16,17,19–21^ are inactive in household, leisure, work, and outdoor activities at 1 year (Supplemental Tables S4 and S5). Whether limitations in I/ADLs (ADLs and IADLs) plateau^10,13,21,28,29^ or increase^11,12,19^ in subsequent years depends on several factors, including age^11,12,29,30^, type of health insurance^11^, and severity of disability 1 to 2 years after stroke^12^.

The level of physical activity has been suggested as one of the factors influencing functional limitations after stroke^31^. Regarding ADLs, some studies have found an association between higher pre-stroke physical activity and lower post-stroke limitations in ADLs^22,32–37^. Specifically, higher pre-stroke physical activity was associated with higher independence in ADLs during the first^22, 32–36^ and second year^37^ after stroke. However, other studies found no evidence of this association between physical activity and functional independence in ADLs^38–41^. These mixed results could be explained by the use of a single-item rating scale^22,32,33,35–41^, the Modified Rankin Scale, which has been shown to be less reliable and more subjective than questionnaires assessing specific I/ADLs^42^. In addition, only one prospective study has examined the effect of pre-stroke physical activity on IADLs^30^. This study focused on vigorous physical activity and was based on a cohort of adults who were stroke-free at baseline. The results showed that higher vigorous physical activity at baseline was associated with a higher probability of being independent in I/ADLs after stroke, but this difference was similar before stroke. This result led the authors to conclude that “being physically active does not protect against the disabling effects of a stroke” on I/ADLs. Building on this previous study, we used a different approach by comparing the effect of physical activity on I/ADLs in a larger sample of stroke survivors (n = 2,143 vs. 1,374) with a sample of stroke-free adults matched for key covariates (n = 10,717). In addition, because moderate-intensity physical activity has been suggested to be at least as beneficial for brain plasticity as vigorous-intensity physical activity^43,44^, both intensities were included in our study.

The objective of this longitudinal case-control study was to examine the effect of pre-stroke physical activity on post-stroke functional limitations. We hypothesized that higher levels of pre-stroke physical activity would reduce post-stroke functional limitations in a cohort of stroke survivors. We further hypothesized that this beneficial effect of physical activity would be weaker in a cohort of stroke-free participants matched for baseline (i.e., before any of the stroke survivors had experienced a stroke) age, sex, body mass index, I/ADL limitations, and country of residence over a similar number of follow-up years, because stroke survivors are likely to have more functional limitations from which to recover.

## METHODS

### Study Sample and Design

Data were drawn from the Survey of Health, Ageing and Retirement in Europe (SHARE), a longitudinal population-based study of over 140,000 adults aged 50 years and older living in 28 European countries and one Middle East country^45^. Data were collected every 2 years between 2004 and 2022 for a total of 8 waves of measurement using computer-assisted personal interviewing (CAPI) in the participants’ homes. Physical activity, stroke events, and functional independence (ADLs, IADLs) were assessed at all measurement waves except wave 3 (2008-2009). To be included in the present study, participants had to be 50 years of age or older, have no reported history of stroke prior to study entry, and have participated in at least 4 waves. SHARE was conducted in accordance with the Declaration of Helsinki and approved by the Ethics Committee of the University of Mannheim (waves 1-4) and the Ethics Council of the Max Planck Society (waves 4-8). All participants gave written informed consent.

### Measures

#### Outcome Variable: Post-Stroke Functional Limitations

Functional dependence was assessed using the number of functional limitations in six ADLs (dressing, walking, bathing, eating, getting in and out of bed, and using the toilet) and seven IADLs (using a map, preparing a hot meal, shopping for groceries, making telephone calls, taking medication, gardening or doing housework, and managing money)^46,47^. Participants were presented with each activity and instructed to indicate whether they “have any difficulty with these activities because of a physical, mental, emotional or memory problem” (yes vs. no) and to “exclude any difficulties they expected to last less than three months”. A score representing the total number of functional dependencies was computed for ADLs (0-6) and IADLs (0-7), with higher scores indicating greater functional dependence.

#### Explanatory Variables: Stroke and Physical Activity

Information on stroke status during follow-up was collected at each wave using the following question: “Has a doctor told you that you have any of the conditions on this card [indicating history of health conditions including stroke]?”^12^.

Level of physical activity at entry in SHARE was derived from two questions: “How often do you engage in vigorous physical activity, such as sports, heavy housework, or a job that involves physical labor?” and “How often do you engage in activities that require a low or moderate level of energy such as gardening, cleaning the car, or doing a walk?”^47–52^. Participants responded on a four-point scale: 1 = Hardly ever or never; 2 = One to three times a month; 3 = Once a week; 4 = More than once a week. Participants who answered “more than once a week” to at least one of the questions were classified as physically active, whereas the other participants were classified as physically inactive to reduce a potential misclassification bias in which physically inactive participants would be incorrectly classified as physically active.

#### Covariates

Models were adjusted for baseline age, sex (male, female), time (survey waves), quadratic time, number of chronic conditions (none or 1 vs. 2 or more), and level of education (primary or less, secondary, tertiary), which has been shown to be associated with physical activity levels^48,51,53–57^.

### Data Preprocessing

#### Matching Procedure

To select matched samples of stroke survivors and stroke-free participants with similar distributions of key covariates, a matching procedure based on the nearest neighbor method was conducted using the MatchIt R package^58,59^ with propensity scores obtained with a generalized linear model. This matching procedure used a 1:5 ratio to create groups including one stroke survivor and five stroke-free adults with similar propensity scores, thereby reducing the potential bias introduced by covariates. Propensity scores were calculated using the characteristics of the participants at their first SHARE interview, when none of them had experienced a stroke: Age, sex, number of chronic conditions (none or 1 vs. 2 or more), limitations in I/ADL, body mass index category [underweight (<18.5 kg/m^2^), normal (reference; 18.5 to 24.9 kg/m^2^), overweight (25 to 29.9 kg/m^2^), obese (30 kg/m^2^ and above)], country of residence, number of measurement waves, and wave number of the first interview.

#### Temporal Alignment of Data on the First Post-Stroke Wave

Using the lag() function of the dplyr R package, which allows the temporal alignment of the measurement waves to be shifted, the first wave in which a stroke was reported was set as Wave 1 in the analyses. Baseline data from stroke survivors used in the matching procedure (see below) were data collected during the wave preceding this Wave 1.

### Statistical Analyses

Data were analyzed using linear mixed-effects models, which account for the nested structure of the data (i.e., repeated measures over time within a single participant), allow the use of incomplete and unbalanced data, and provide acceptable Type I error rates^60,61^. Models were built and fitted by maximum likelihood in R programming language^62^ using the lme4^63^ and lmerTest^64^ packages. P-values were approximated using Satterthwaite method^64^. The effects of baseline physical activity on future I/ADL were compared between stroke survivors and stroke-free adults. The number of post-stroke measurement waves varied between participants in the stroke group and was matched to the number of measurement waves in the stroke-free adults. To examine the effect of pre-stroke physical activity on functional independence in stroke survivors and stroke-free adults, two dependent variables were tested: ADL and IADL limitations. The fitted models included stroke (stroke vs. no stroke), physical activity (active vs. inactive at baseline), linear time, quadratic time, and the covariates as fixed effects. The random structure included random intercepts for participants and for participants grouped together by the matching process, as well as random linear and quadratic slopes of wave within each participant^61^. These random effects estimated the functional independence of each participant and each matching group, as well as the rate of change of this independence across waves. The quadratic effect of wave was added to account for the potential accelerated (or decelerated) decline in functional independence across waves. An interaction term between stroke and physical activity conditions was added to formally test the moderating effect of stroke on the association between physical activity and functional dependence. In addition, interaction terms between stroke and wave (linear and quadratic) conditions were included to allow variations in I/ADL trajectories between the two groups. In summary, the equation of our models was as follows:

Functional Limitation_ij_

= *β*_0_ + *β*_1_ Stroke Status_j_ + Baseline Physical Activity_j_

+ *β*_3_ (Stroke Status_j_ × Baseline Physical Activity_j_) + *β*_4_ Wave_ij_

+ *β*_5_ (Stroke Status_j_ × Wave_ij_) + *β*_6_ Quadratic Wave_ij_

+ *β*_7_ (Stroke Status_j_ × Quadratic Wave_ij_) + Baseline Age_j_ + *β*_9_ Sex_j_

+ *β*_10_ Education Primary_j_ + *β*_11_ Education Tertiary_j_

+ *β*_12_ Chronic Health Conditions_j_ + *u*_0j_ + *u*_k(j)_ + *u*_1j_ × *Wave*_ij_

+ *u*_2j_ × *Quadratic Wave*_ij_ + *∈_ij_*

In this equation, *Functional Limitationij* is the j^th^ participant’s number of limitations in ADLs or IADLs in condition *i*, the *β*s are the fixed effect coefficients, *u*_0j_ is the random intercept for the *j*^th^ participant, *u*_1j_ is the random slope of the wave for the *j*^th^ participant, *u*_2j_ is the random slope of the quadratic wave for the *j*^th^ participant, *u*_k(j)_ is a random intercept with *k(j)* coding for the matching group of the *j*^th^ participant, the random effects are allowed to correlate freely (unstructured covariance matrix), and *∈_ij_* is the error term.

#### Sensitivity Analysis

In a sensitivity analysis, participants who responded “hardly ever or never” to one of the two physical activity questions were classified as physically inactive, whereas the other participants were classified as physically active. This categorization reduced a potential misclassification bias in which physically active participants would be incorrectly classified as physically inactive.

## RESULTS

### Descriptive Results

The study sample included 2,143 stroke survivors (mean age: 66.9 ± 9.1 years; 1,052 females) and 10,717 stroke-free adults (mean age: 66.9 ± 9.3 years, 5,126 females) from the SHARE study, whose characteristics at baseline are summarized in Table 1.

**Table 1.** Baseline characteristics of the participants at their first interview for the Survey of Health, Ageing and Retirement in Europe (SHARE), when none of them had experienced a stroke, stratified by stroke-related status in the following waves.

| Variables | Stroke Survivors<br>(N = 2,143) | Matched Stroke-Free Adults<br>(N = 10,717) |
| --- | --- | --- |
| **Age**, mean (SD) | 66.9 (9.1) | 66.9 (9.3) |
| **Sex** |  |  |
| Female, n (%) | 1052 (49.1) | 5126 (47.8) |
| Male, n (%) | 1091 (50.9) | 5591 (52.2) |
| **Physical Activity** |  |  |
| Physically active, n (%) | 1564 (72.5) | 8060 (74.6) |
| Physically inactive, n (%) | 595 (27.4) | 2720 (25.2) |
| **Functional Limitations** |  |  |
| ADL, mean (SD) | 0.2 (0.6) | 0.2 (0.7) |
| IADL, mean (SD) | 0.3 (0.8) | 0.3 (0.9) |
| **Body Mass Index (kg/m<sup>2</sup>)** |  |  |
| < 18.5 – Underweight, n (%) | 155 (1.5) | 590 (1.1) |
| 18.5-24.9 – Normal, n (%) | 3445 (33.9) | 16275 (31.7) |
| 25-29.9 – Overweight, n (%) | 4176 (41.0) | 22856 (44.5) |
| ≥ 30 – Obese, n (%) | 2401 (23.6) | 11647 (22.7) |
| **Chronic Condition** |  |  |
| < 2, n (%) | 3423 (32.7) | 22807 (43.5) |
| ≥ 2, n (%) | 7053 (67.3) | 29676 (56.5) |
| **Education** |  |  |
| Primary, n (%) | 666 (31.1) | 3027 (28.2) |
| Secondary, n (%) | 1081 (50.4) | 5415 (50.5) |
| Tertiary, n (%) | 396 (18.5) | 2275 (21.2) |
| **Country** |  |  |
| Austria, n (%) | 147 (6.9) | 764 (7.1) |
| Belgium, n (%) | 193 (9.0) | 965 (9.0) |
| Czech Republic, n (%) | 160 (7.5) | 818 (7.6) |
| Denmark, n (%) | 170 (7.9) | 810 (7.6) |
| Estonia, n (%) | 154 (7.2) | 870 (8.1) |
| France, n (%) | 161 (7.5) | 815 (7.6) |
| Germany, n (%) | 153 (7.1) | 831 (7.8) |
| Greece, n (%) | 106 (4.9) | 506 (4.7) |
| Israel, n (%) | 95 (4.4) | 460 (4.3) |
| Italy, n (%) | 161 (7.5) | 768 (7.2) |
| Luxembourg, n (%) | 20 (0.9) | 94 (0.9) |
| Netherlands, n (%) | 81 (3.8) | 384 (3.6) |
| Poland, n (%) | 73 (3.4) | 368 (3.4) |
| Slovenia, n (%) | 76 (3.5) | 379 (3.5) |
| Spain, n (%) | 141 (6.6) | 673 (6.3) |
| Sweden, n (%) | 167 (7.8) | 812 (7.6) |
| Switzerland, n (%) | 85 (4.0) | 404 (3.8) |

Notes. ADL = activities of daily living, IADL = instrumental activities of daily living, SD = standard deviation.

In stroke survivors, the average level of functional limitation at the first wave after stroke was 0.17 for ADLs and 0.28 for IADLs and increased to 1.05 and 1.72 at the eighth wave after stroke, respectively (Supplemental Table S6). In stroke-free adults, the average level of functional limitation on the first wave after stroke was 0.15 for ADLs and 0.25 for IADLs and increased to 0.41 and 0.73 on the eighth wave after stroke, respectively (Supplemental Table S6). The similar level of functional limitation between groups at the first poststroke wave was expected because the propensity score used for the matching procedure included the number of I/ADL limitations at baseline.

At baseline, 258 stroke survivors engaged in moderate physical activity “hardly ever or never”, 127 “one to three times a month”, 274 “once a week”, and 1,500 “more than once a week” (Supplemental Table S7). In addition, 951 engaged in vigorous physical activity “hardly ever or never”, 216 “one to three times a month”, 306 “once a week”, and 706 “more than once a week” (Supplemental Table S7). In total, 1,564 stroke survivors were considered physically active (i.e., answered “more than once a week” to at least one of the two questions) and 595 were considered physically inactive (Supplemental Table S8).

### Statistical Results

Results of the mixed-effects models showed an interaction effect between stroke status and physical activity on ADL limitations (b = -0.076, 95% confidence interval [CI]: -0.142 to -0.011, p = 0.022; Table 2, Figure 1). The simple effects of the terms in this interaction confirmed that the effect of physical activity was stronger in stroke survivors (b = -0.345, 95% CI: -0.438, -0.252, p < 2.0 × 10^-16^) than in stroke-free adults (b = -0.269, 95% CI: -0.296 to -0.241, p < 2.0 × 10^-16^), with physically active participants (i.e., physical activity > once a week) showing fewer limitations in ADLs than physically inactive participants (i.e., physical activity ≤ once a week). Similarly, a main effect showed that physically active participants showed fewer limitations in IADLs (b = - 0.410, 95% CI: -0.445 to -0.375, p < 2.0 × 10^-16^; Table 2, Figure 1). However, results showed no evidence of an interaction effect of stroke status and physical activity on limitations in IADLs (b = -0.057, 95% CI: -0.140 to 0.026, p = 0.178; Table 2, Figure 1).

**Figure 1.**
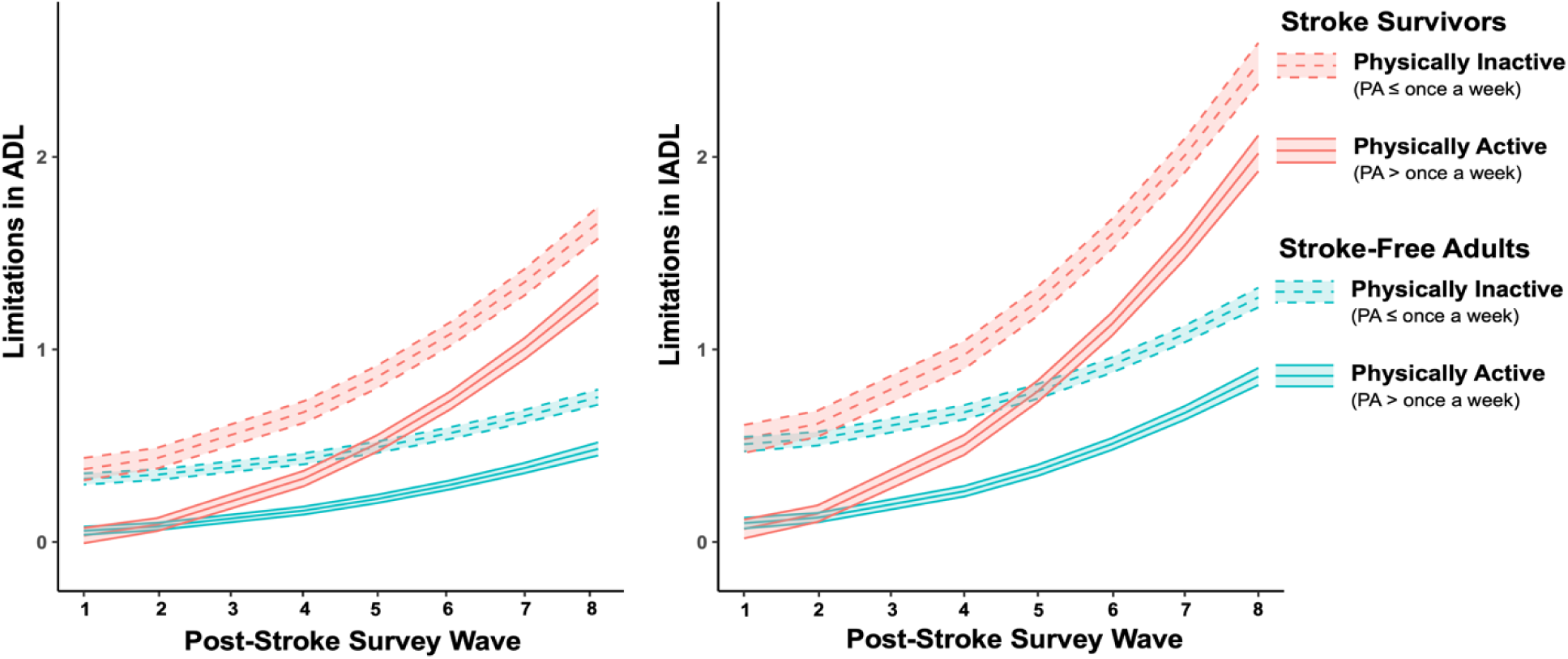
Effect of physical activity (PA ≤ once a week vs. PA > once a week) on limitations in activities of daily living (ADL, left panel) and instrumental activities of daily living (IADL, right panel) in stroke survivors and matched stroke-free adults.

**Table 2.** Results of the mixed-effects models testing the interaction between stroke-related status and physical activity (PA; once a week or less vs. more than once a week) on limitations in activities of daily living (ADL) and instrumental activities of daily living (IADL).

| Exposures | ADL |  | IADL |  |
| --- | --- | --- | --- | --- |
|  | b (95 CI) | p | b (95 CI) | p |
| Intercept | -0.379 (-0.472 to -0.286) | $1.3 \times 10^{-15}$ | -0.849 (-0.971 to -0.727) | $< 2.0 \times 10^{-16}$ |
| Stroke | 0.051 (-0.011 to 0.113) | 0.105 | 0.027 (-0.050 to 0.104) | 0.495 |
| Physical Activity | -0.269 (-0.296 to -0.241) | $< 2.0 \times 10^{-16}$ | -0.410 (-0.445 to -0.375) | $< 2.0 \times 10^{-16}$ |
| Wave | 0.016 (0.005 to 0.027) | 0.005 | 0.015 (0.001 to 0.029) | 0.030 |
| Wave<sup>2</sup> | 0.006 (0.005 to 0.008) | $5.7 \times 10^{-15}$ | 0.013 (0.011 to 0.015) | $< 2.0 \times 10^{-16}$ |
| Age | 0.008 (0.007 to 0.009) | $< 2.0 \times 10^{-16}$ | 0.014 (0.013 to 0.016) | $< 2.0 \times 10^{-16}$ |
| Sex | 0.073 (0.051 to 0.095) | $1.1 \times 10^{-10}$ | 0.204 (0.175 to 0.232) | $< 2.0 \times 10^{-16}$ |
| Education |  |  |  |  |
| Primary (vs. Secondary) | 0.115 (0.090 to 0.141) | $< 2.0 \times 10^{-16}$ | 0.219 (0.187 to 0.251) | $< 2.0 \times 10^{-16}$ |
| Tertiary (vs. Secondary) | -0.024 (-0.053 to 0.004) | 0.094 | -0.043 (-0.079 to -0.007) | 0.020 |
| Chronic Conditions | 0.115 (0.101 to 0.130) | $< 2.0 \times 10^{-16}$ | 0.180 (0.162 to 0.198) | $< 2.0 \times 10^{-16}$ |
| Stroke × Physical Activity | -0.076 (-0.142 to -0.011) | 0.022 | -0.057 (-0.140 to 0.026) | 0.178 |
| Stroke × Wave | 0.020 (-0.006 to 0.047) | 0.128 | 0.031 (-0.002 to 0.064) | 0.066 |
| Stroke × Wave<sup>2</sup> | 0.015 (0.011 to 0.018) | $4.9 \times 10^{-13}$ | 0.020 (0.015 to 0.025) | $3.9 \times 10^{-15}$ |

Notes. 95 CI = 95% confidence interval, ADL = activities of daily living, IADL = instrumental activities of daily living

An interaction between stroke status and quadratic wave indicated that limitations in ADLs (b = 0.015, 95% CI: 0.011 to 0.018, p = 4.9 × 10^-13^) and IADLs (b = 0.020, 95% CI: 0.015 to 0.025, p = 3.9 × 10^-15^) increased across waves at a higher rate in stroke survivors than in stroke-free adults (Figure 1).

Results of the sensitivity analyses based on a different threshold for classifying physically active (i.e., physical activity ≥ once a month) and inactive participants (i.e., hardly ever or never active) were consistent with the results of the main analyses (Supplemental Table S9; Supplemental Figure S1).

## DISCUSSION

### Main Results

The objective of this longitudinal case-control study was to examine the effect of pre-stroke physical activity on post-stroke functional limitations in stroke survivors and matched stroke-free adults. Consistent with our hypothesis, the results suggest that the beneficial effect of pre-stroke physical activity on post-stroke ADL limitations is stronger than its effect in stroke-free adults matched for age, sex, body mass index, limitations in I/ADLs, number of chronic conditions, and country of residence, number of measurement waves, and wave number of the first interview.

### Comparison With Other Studies

Our results in stroke survivors showed that higher levels of pre-stroke physical activity were associated with fewer ADL limitations, which is consistent with the existing literature^22,32–37^. Most importantly, our results reveal that the effect of pre-stroke physical activity on in ADL limitations after stroke is statistically stronger than its effect in matched stroke-free adults. Although the study by Ris et al.^30^ also examined the effect of physical activity in both stroke survivors and stroke-free adults (without the matching procedure we conducted), this potential interaction effect was not considered. Several mechanisms could explain how physical activity improves functional independence after stroke, such as an association between pre- and post-stroke physical activity. Previous studies have shown that this level of physical activity is similar in 41 to 42% of stroke survivors activity^66,67^. Post-stroke engagement in physical activity could increase brain plasticity processes such as angiogenesis, synaptogenesis, and neurogenesis, primarily through the upregulation of growth factors (e.g., brain-derived neurotrophic factor; BDNF)^68–70^. However, the same studies also showed that 33 to 39% of stroke survivors reported lower levels of physical activity after stroke compared to before stroke, and 20 to 25% reported higher levels of physical activity^66,67^. Another explanation could be the beneficial effect of pre-stroke physical activity on depression^66^, which has been shown to be associated with ADL limitations^47,71,72^.

### Strengths and Limitations

The present study has several strengths including results presented for a long follow-up period (up to 16 years) and a large international post-stroke population (17 countries), which allowed us to robustly examine the effects of physical activity on I/ADL limitations. The number of I/ADL limitations was used to assess changes in functional limitation over time, which is more reliable than single-item ratings and more sensitive to identifying differences in functional trajectories between stroke survivors and stroke-free adults. Sensitivity results using a different categorization of physical activity were consistent with the main results.

However, our results should be considered in the light of several limitations. (1) There was a lack of information on stroke characteristics, which is common in and inherent to large-scale longitudinal studies. Differences in stroke subtypes (i.e., ischemic, hemorrhagic, cryptogenic, transient ischemic attack) or in the type of impairment resulting from the stroke (e.g., motor, sensory, visual, cognitive) may partly explain the discrepancy in our findings between ADL and IADL. The different behaviors that comprise ADL (which rely more on basic motor functions) and IADL (which rely more on cognitive functions) may interact with stroke characteristics. However, data specifying stroke characteristics is not available in SHARE. Future studies should be supported by medical records to provide a more specific understanding of the relationship between physical activity and specific aspects of functional independence by stroke characteristics. (2) The outcome (i.e., stroke) was self-reported. Therefore, a memory bias cannot be excluded. However, the agreement between self-reported stroke and medical records ranges from 79%^73^ to 96%^74^. (3) Physical activity was self-reported, which may not have accurately captured the actual levels of physical activity, as correlations between self-report and direct measures of physical activity are low to moderate^75,76^. Future studies should assess physical activity using device-based measures, which have been shown to have greater validity and reliability^77^.

### Conclusions

Our results support a stronger long-term beneficial effect of physical activity on independence in ADLs in stroke survivors compared with stroke-free adults. These findings underscore the beneficial role of moderate-to-vigorous physical activity in mitigating stroke-related limitations in ADLs. In addition, these findings highlight the need to consider the pre-stroke levels of physical activity in the prognosis of stroke-related functional independence.

As movement specialists and primary care practitioners, physical therapists are key healthcare professionals in the prevention of physical inactivity, which falls within their scope of practice^78,79^. As such, the expertise of physical therapists should be used to help people achieve the recommendations of physical activity, thereby optimizing their functional independence in the event of a stroke. While physical therapists feel confident in providing general advice to patients and clients about a physically active lifestyle and suggesting specific physical activity programs, they also perceive some barriers in providing this comprehensive care, including the lack of time, counseling skills, and reimbursement^80^. Such reimbursement may lead to the emergence of certified clinical specialists who can develop mor in-depth knowledge and skills related to physical inactivity.

## Data Availability

The SHARE dataset is available at http://www.share-project.org/data-access.html and the DOIs for the waves used in the current study are the following: https://doi.org/10.6103/SHARE.w1.600, https://doi.org/10.6103/SHARE.w2.600, https://doi.org/10.6103/SHARE.w4.600, https://doi.org/10.6103/SHARE.w5.600, https://doi.org/10.6103/SHARE.w6.600, https://doi.org/10.6103/SHARE.w7.711, https://doi.org/10.6103/SHARE.w8cabeta.001.

https://github.com/matthieuboisgontier/Stroke_Physical-Activity

## Nonstandard abbreviations and acronyms

ADL: activities of daily living
IADL: instrumental activities of daily living
SHARE: Survey of Health, Ageing and Retirement in Europe.

## ARTICLE INFORMATION

## Acknowledgements

Based on the Contributor Roles Taxonomy (CRediT)^81,82^, individual author contributions to this work are as follows: Zack van Allen: Formal Analysis; Visualization; Data Curation; Writing – Review and Editing; Dan Orsholits: Methodology; Formal Analysis; Visualization; Data Curation; Writing – Review and Editing; Matthieu P. Boisgontier: Conceptualization; Methodology; Formal Analysis; Visualization; Data Curation; Writing – Original Draft; Writing – Review and Editing; Supervision; Project Administration; Funding Acquisition.

## Funding

Dr Boisgontier is supported by the Canada Foundation for Innovation (CFI), Mitacs, and the Banting Research Foundation. Zack van Allen is supported by a Mitacs-Banting Discovery Postdoctoral Fellowship. The SHARE data collection was primarily funded by the European Commission through FP5 (QLK6-CT-2001-00360), FP6 (SHARE-I3: RII-CT-2006-062193, COMPARE: CIT5-CT-2005-028857, SHARELIFE: CIT4-CT-2006-028812) and FP7 (SHARE-PREP: no.211909, SHARE-LEAP: no.227822, SHARE M4: no.261982). Additional funding from the German Ministry of Education and Research, the Max Planck Society for the Advancement of Science, the U.S. National Institute on Aging (U01_AG09740-13S2, P01_AG005842, P01_AG08291, P30_AG12815, R21_AG025169, Y1-AG-4553-01, IAG_BSR06-11, OGHA_04-064, HHSN271201300071C) and from various national funding sources is gratefully acknowledged (see www.share-project.org).

## Data and Code Sharing

In accordance with good research practices^83^, the R code used to analyze the data is publicly available online^84^. A non-peer-reviewed version of this manuscript is publicly available online^85^. The SHARE dataset is available at http://www.share-project.org/data-access.html and the DOIs for the waves used in the current study are the following: https://doi.org/10.6103/SHARE.w1.600, https://doi.org/10.6103/SHARE.w2.600, https://doi.org/10.6103/SHARE.w4.600, https://doi.org/10.6103/SHARE.w5.600, https://doi.org/10.6103/SHARE.w6.600, https://doi.org/10.6103/SHARE.w7.711, https://doi.org/10.6103/SHARE.w8cabeta.001.

## Reporting Guidelines

This manuscript conforms to the STROBE guidelines for observational studies (http://www.strobe-statement.org)^86^.

## Acknowledgements

The authors are grateful to Dr. Olivier Renaud (University of Geneva, Switzerland) for his help in transcribing the R code of our statistical models into a scientific equation.

## SUPPLEMENTAL MATERIAL

**Table S1.**
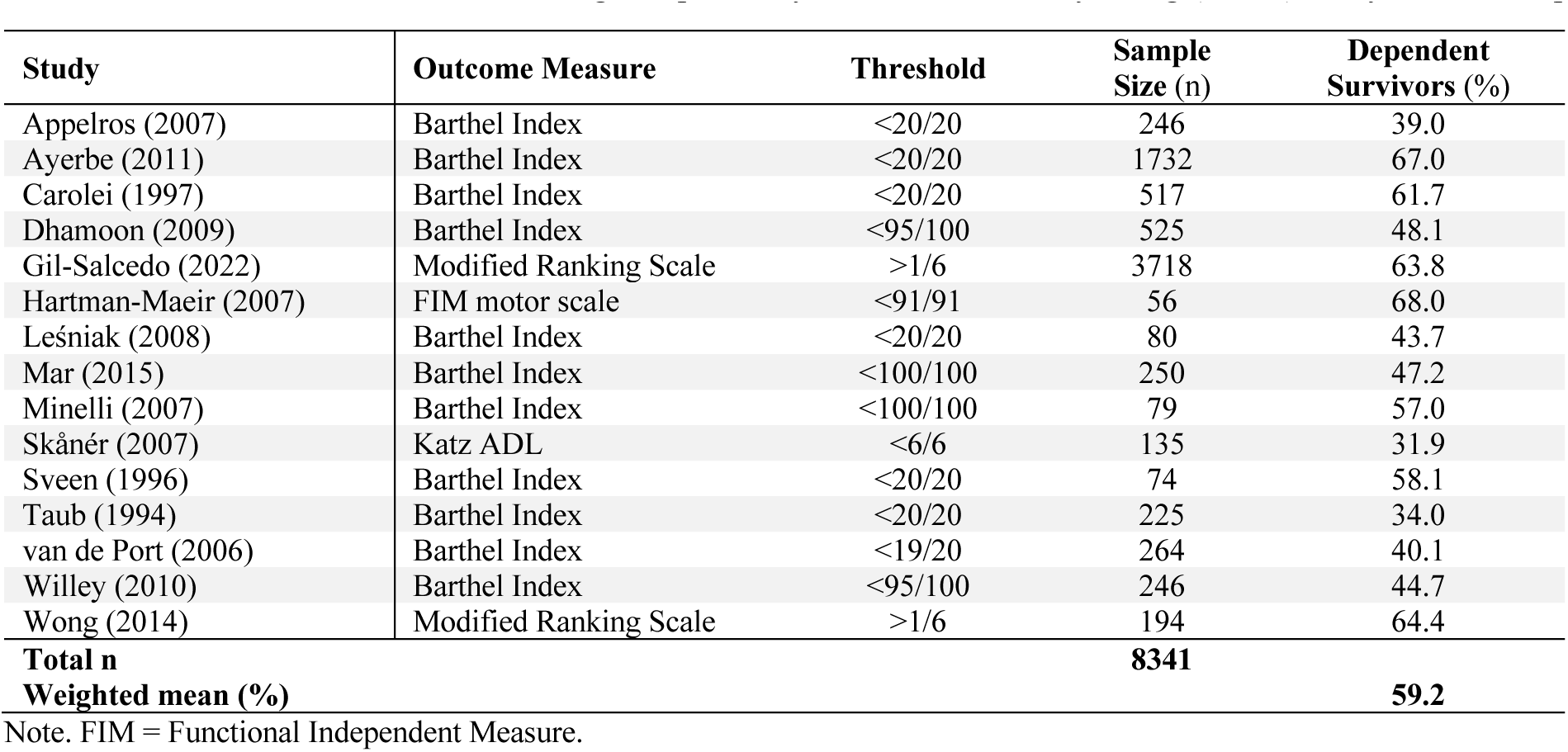
Stroke survivors with at least slight dependency in activities of daily living (ADLs) at 1-year follow-up.

**Table S2.** Stroke survivors with at least moderate dependency in activities of daily living (ADLs) at 1-year follow-up.

| Study | Outcome Measure | Threshold | Sample Size (n) | Dependent Survivors (%) |
| --- | --- | --- | --- | --- |
| Appelros (2007) | Barthel Index | <15/20 | 246 | 31.2 |
| Broussy (2019) | Modified Ranking Scale | >2/6 | 161 | 29.6 |
| De Campos (2017) | Modified Ranking Scale | >2/6 | 287 | 16.4 |
| Jokinen (2015) | Modified Ranking Scale | >2/6 | 364 | 44.0 |
| López-Cancio (2017) | Modified Ranking Scale | >2/6 | 143 | 53.8 |
| Mar (2015) | Barthel Index | <90/100 | 250 | 40.4 |
| Patel (2002) | Barthel Index | <15/20 | 619 | 36.2 |
| Patel (2003) | Barthel Index | <15/20 | 136 | 36.0 |
| Santus (1990) | Barthel Index | <75/100 | 76 | 46.1 |
| Taub (1994) | Barthel Index | <15/20 | 225 | 11.0 |
| Urbanek (2018) | Modified Ranking Scale | >2/6 | 1119 | 41.6 |
| Verhoeven (2011) | Barthel Index | <18/20 | 92 | 38.0 |
| Wafa (2020) | Barthel Index | <15/20 | 1961 | 24.1 |
| Wolfe (2011) | Barthel Index | <15/20 | 1578 | 13.1 |
| Wong (2014) | Modified Ranking Scale | >2/6 | 194 | 33.0 |
| **Total (n)** |  |  | **7451** |  |
| **Weighted mean (%)** |  |  |  | **32.9** |

**Table S3.** Stroke survivors with severe or total dependency in activities of daily living (ADLs) at 1-year follow-up.

| Study | Outcome Measure | Threshold | Sample Size (n) | Dependent Survivors (%) |
| --- | --- | --- | --- | --- |
| Appelros (2007) | Barthel Index | <12/20 | 246 | 16.0 |
| Broussy (2019) | Barthel Index | <12/20 | 161 | 12.7 |
| Dhamoon (2009) | Barthel Index | <60/100 | 525 | 18.0 |
| Gil-Salcedo (2022) | Modified Ranking Scale | >3/6 | 3718 | 27.3 |
| Mar (2015) | Barthel Index | <60/100 | 250 | 20.4 |
| Patel (2002) | Barthel Index | <10/20 | 619 | 9.4 |
| Patel (2003) | Barthel Index | <10/20 | 136 | 15.4 |
| Wiley (2010) | Barthel Index | <60/100 | 246 | 15.9 |
| Wong (2014) | Modified Ranking Scale | >3/6 | 194 | 19.6 |
| **Total n** |  |  | **6095** |  |
| **Weighted mean (%)** |  |  |  | **22.6** |

**Table S4.** Stroke survivors who are moderately active in instrumental activities of daily living (IADLs) at 1-year follow-up.

| Study | Outcome Measure | Threshold | Sample Size (n) | Dependent Survivors (%) |
| --- | --- | --- | --- | --- |
| Appelros (2007) | Frenchay Activities Index | <30/45 | 246 | 78.8 |
| Ayerbe (2011) | Frenchay Activities Index | <30/45 | 1403 | 79.7 |
| Patel (2002) | Frenchay Activities Index | <30/45 | 619 | 85.7 |
| Patel (2003) | Frenchay Activities Index | <30/45 | 136 | 88.2 |
| Sveen (1996) | Frenchay Activities Index | <29/45 | 74 | 75.6 |
| **Total n** |  |  | **2478** |  |
| **Weighted mean (%)** |  |  |  | **81.5** |

**Table S5.** Stroke survivors who are inactive in instrumental activities of daily living (IADLs) at 1-year follow-up.

| Study | Outcome Measure | Threshold | Sample Size (n) | Dependent Survivors (%) |
| --- | --- | --- | --- | --- |
| Appelros (2007) | Frenchay Activities Index | <15/45 | 246 | 46.3 |
| Patel (2002) | Frenchay Activities Index | <15/45 | 619 | 40.4 |
| Patel (2003) | Frenchay Activities Index | <15/45 | 136 | 72.7 |
| van de Port (2006) | Frenchay Activities Index | <15/45 | 264 | 35.2 |
| Wolfe (2011) | Frenchay Activities Index | <15/45 | 1578 | 38.8 |
| **Total n** |  |  | **2843** |  |
| **Weighted mean (%)** |  |  |  | **41.1** |

**Table S6.**
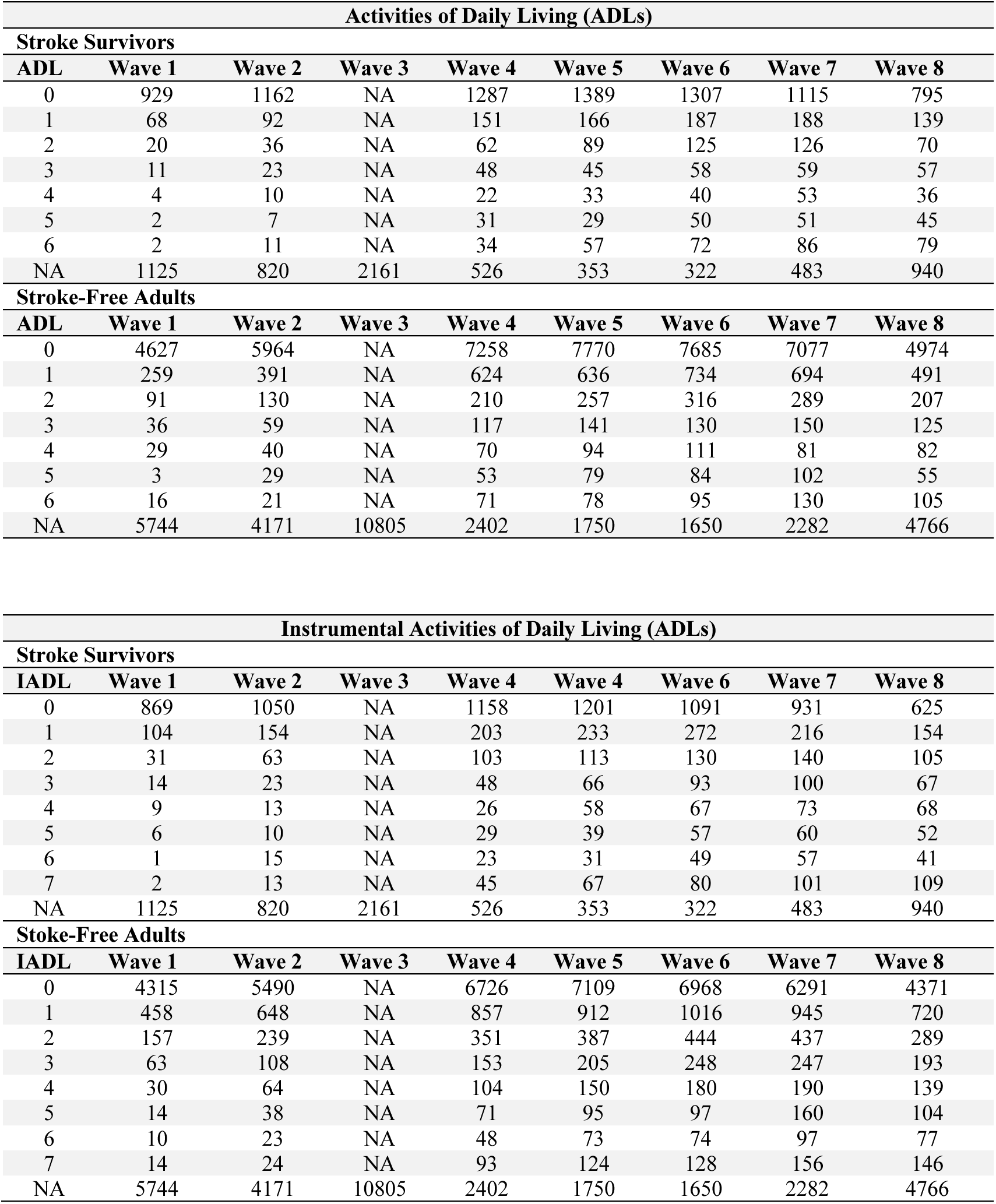
Activities of daily living (ADLs) and instrumental activities of daily living (IADLs) counts for stroke survivors and stroke-free adults. Counts reflect the number of I/ADL limitations reported.

| Activities of Daily Living (ADLs) |  |  |  |  |  |  |  |  |
| --- | --- | --- | --- | --- | --- | --- | --- | --- |
| Stroke Survivors |  |  |  |  |  |  |  |  |
| ADL | Wave 1 | Wave 2 | Wave 3 | Wave 4 | Wave 5 | Wave 6 | Wave 7 | Wave 8 |
| 0 | 929 | 1162 | NA | 1287 | 1389 | 1307 | 1115 | 795 |
| 1 | 68 | 92 | NA | 151 | 166 | 187 | 188 | 139 |
| 2 | 20 | 36 | NA | 62 | 89 | 125 | 126 | 70 |
| 3 | 11 | 23 | NA | 48 | 45 | 58 | 59 | 57 |
| 4 | 4 | 10 | NA | 22 | 33 | 40 | 53 | 36 |
| 5 | 2 | 7 | NA | 31 | 29 | 50 | 51 | 45 |
| 6 | 2 | 11 | NA | 34 | 57 | 72 | 86 | 79 |
| NA | 1125 | 820 | 2161 | 526 | 353 | 322 | 483 | 940 |
| Stroke-Free Adults |  |  |  |  |  |  |  |  |
| ADL | Wave 1 | Wave 2 | Wave 3 | Wave 4 | Wave 5 | Wave 6 | Wave 7 | Wave 8 |
| 0 | 4627 | 5964 | NA | 7258 | 7770 | 7685 | 7077 | 4974 |
| 1 | 259 | 391 | NA | 624 | 636 | 734 | 694 | 491 |
| 2 | 91 | 130 | NA | 210 | 257 | 316 | 289 | 207 |
| 3 | 36 | 59 | NA | 117 | 141 | 130 | 150 | 125 |
| 4 | 29 | 40 | NA | 70 | 94 | 111 | 81 | 82 |
| 5 | 3 | 29 | NA | 53 | 79 | 84 | 102 | 55 |
| 6 | 16 | 21 | NA | 71 | 78 | 95 | 130 | 105 |
| NA | 5744 | 4171 | 10805 | 2402 | 1750 | 1650 | 2282 | 4766 |

| Instrumental Activities of Daily Living (ADLs) |  |  |  |  |  |  |  |  |
| --- | --- | --- | --- | --- | --- | --- | --- | --- |
| Stroke Survivors |  |  |  |  |  |  |  |  |
| IADL | Wave 1 | Wave 2 | Wave 3 | Wave 4 | Wave 4 | Wave 6 | Wave 7 | Wave 8 |
| 0 | 869 | 1050 | NA | 1158 | 1201 | 1091 | 931 | 625 |
| 1 | 104 | 154 | NA | 203 | 233 | 272 | 216 | 154 |
| 2 | 31 | 63 | NA | 103 | 113 | 130 | 140 | 105 |
| 3 | 14 | 23 | NA | 48 | 66 | 93 | 100 | 67 |
| 4 | 9 | 13 | NA | 26 | 58 | 67 | 73 | 68 |
| 5 | 6 | 10 | NA | 29 | 39 | 57 | 60 | 52 |
| 6 | 1 | 15 | NA | 23 | 31 | 49 | 57 | 41 |
| 7 | 2 | 13 | NA | 45 | 67 | 80 | 101 | 109 |
| NA | 1125 | 820 | 2161 | 526 | 353 | 322 | 483 | 940 |
| Stroke-Free Adults |  |  |  |  |  |  |  |  |
| IADL | Wave 1 | Wave 2 | Wave 3 | Wave 4 | Wave 5 | Wave 6 | Wave 7 | Wave 8 |
| 0 | 4315 | 5490 | NA | 6726 | 7109 | 6968 | 6291 | 4371 |
| 1 | 458 | 648 | NA | 857 | 912 | 1016 | 945 | 720 |
| 2 | 157 | 239 | NA | 351 | 387 | 444 | 437 | 289 |
| 3 | 63 | 108 | NA | 153 | 205 | 248 | 247 | 193 |
| 4 | 30 | 64 | NA | 104 | 150 | 180 | 190 | 139 |
| 5 | 14 | 38 | NA | 71 | 95 | 97 | 160 | 104 |
| 6 | 10 | 23 | NA | 48 | 73 | 74 | 97 | 77 |
| 7 | 14 | 24 | NA | 93 | 124 | 128 | 156 | 146 |
| NA | 5744 | 4171 | 10805 | 2402 | 1750 | 1650 | 2282 | 4766 |

**Table S7.** Frequency of moderate and vigorous physical activity in stroke survivors and stroke-free adults at baseline.

| Physical Activity Frequency | Stroke Survivors |  | Stroke-Free Adults |  |
| --- | --- | --- | --- | --- |
|  | Moderate Physical Activity | Vigorous Physical Activity | Moderate Physical Activity | Vigorous Physical Activity |
| Hardly ever or never | 258 | 931 | 1086 | 4291 |
| One to three times a month | 127 | 216 | 575 | 1050 |
| Once a week | 274 | 306 | 1415 | 1501 |
| More than once a week | 1500 | 706 | 7702 | 3937 |
| NA | 2 | 2 | 27 | 26 |

Notes. The question for assessing moderate physical activity was “How often do you engage in activities that require a low or moderate level of energy such as gardening, cleaning the car, or doing a walk?”. The question for assessing vigorous physical activity was “How often do you engage in vigorous physical activity, such as sports, heavy housework, or a job that involves physical labor?”

**Table S8.** Counts of physically active and inactive participants in stroke survivors and stroke-free adults.

| Type of Analysis | Stroke Survivors |  | Stroke-Free Adults |  |
| --- | --- | --- | --- | --- |
|  | Physically Inactive | Physically Active | Physically Inactive | Physically Active |
| Main Analyses | 595 | 1564 | 2720 | 8060 |
| Sensitivity Analyses | 1202 | 957 | 6343 | 4434 |

Notes. In the main analyses, participants who answered “more than once a week” to at least one of the questions were classified as physically active, whereas the other participants were classified as physically inactive. In the sensitivity analyses, participants who answered “hardly ever or never” to one of the two questions related to the level of physical activity were classified as physically inactive, whereas the other participants were classified as physically active.

**Table S9.** Results of the sensitivity analyses testing the interaction between stroke-related status and physical activity (“hardly ever or never” vs. “at least once a month”) on limitations in activities of daily living (ADLs) and instrumental activities of daily living (IADLs).

| Exposures | ADL |  | IADL |  |
| --- | --- | --- | --- | --- |
|  | b (95 CI) | p | b (95 CI) | p |
| Intercept | -0.563 (-0.653 to -0.474) | $< 2.0 \times 10^{-16}$ | -1.137 (-1.255 to -1.019) | $< 2.0 \times 10^{-16}$ |
| Stroke | -0.044 (-0.089 to 0.001) | 0.056 | -0.045 (-0.100 to 0.010) | 0.107 |
| Physical Activity | 0.223 (0.198 to 0.247) | $< 2.0 \times 10^{-16}$ | 0.317 (0.285 to 0.348) | $< 2.0 \times 10^{-16}$ |
| Wave | 0.016 (0.005 to 0.027) | 0.003 | 0.017 (0.003 to 0.030) | 0.015 |
| Wave<sup>2</sup> | 0.006 (0.005 to 0.008) | $1.1 \times 10^{-14}$ | 0.013 (0.011 to 0.015) | $< 2.0 \times 10^{-16}$ |
| Age | 0.007 (0.005 to 0.008) | $< 2.0 \times 10^{-16}$ | 0.012 (0.011 to 0.014) | $< 2.0 \times 10^{-16}$ |
| Sex | 0.063 (0.041 to 0.085) | $3.0 \times 10^{-8}$ | 0.192 (0.164 to 0.221) | $< 2.0 \times 10^{-16}$ |
| Education |  |  |  |  |
| Primary (vs. Secondary) | 0.117 (0.091 to 0.142) | $< 2.0 \times 10^{-16}$ | 0.224 (0.191 to 0.256) | $< 2.0 \times 10^{-16}$ |
| Tertiary (vs. Secondary) | -0.026 (-0.055 to 0.003) | 0.074 | -0.047 (-0.083 to -0.011) | 0.011 |
| Chronic Conditions | 0.113 (0.099 to 0.128) | $< 2.0 \times 10^{-16}$ | 0.178 (0.160 to 0.196) | $< 2.0 \times 10^{-16}$ |
| Stroke × Physical Activity | 0.084 (0.026 to 0.142) | 0.004 | 0.062 (-0.012 to 0.136) | 0.098 |
| Stroke × Wave | 0.022 (-0.005 to 0.048) | 0.109 | 0.032 (0.000 to 0.065) | 0.052 |
| Stroke × Wave<sup>2</sup> | 0.014 (0.010 to 0.018) | $7.9 \times 10^{-13}$ | 0.020 (0.015 to 0.025) | $6.7 \times 10^{-15}$ |

Notes. 95 CI = 95% confidence interval, ADL = activities of daily living, IADL = instrumental activities of daily living

**Figure S1.**
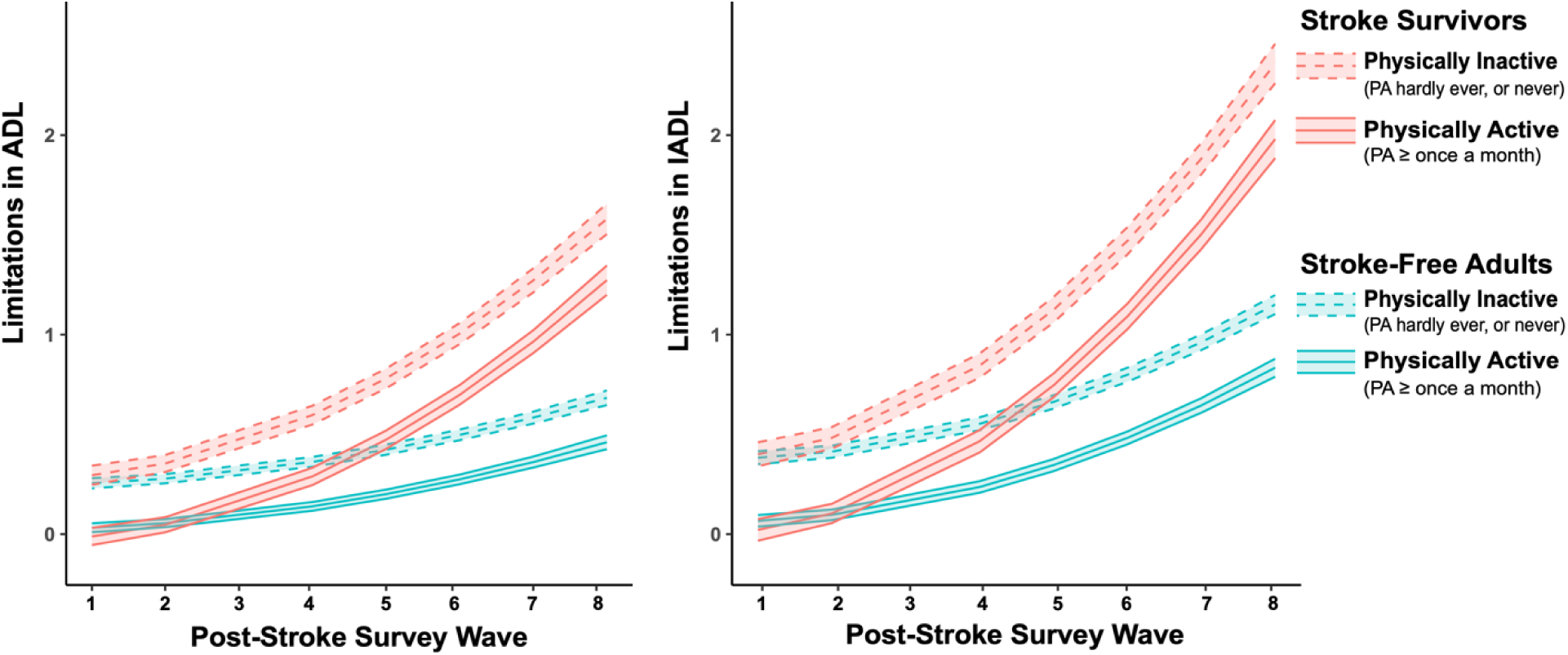
Result of the sensitivity analysis showing the effect of physical activity (“Hardly ever or never” vs. “At least once a month”) on limitations in activities of daily living (ADLs) and instrumental activities of daily living (IADLs) in stroke survivors and matched stroke-free adults over time.

## Notes

### Competing Interest Statement

The authors have declared no competing interest.

### Author Declarations

SHARE was carried out in accordance with the Declaration of Helsinki and has been approved by the Ethics Committee of the University of Mannheim (waves 1-4) and the Ethics Council of the Max Plank Society (waves 4-8). All participants provided written informed consent. The University of Ottawa Office of Research Ethics and Integrity formally confirmed that the study did not require Research Ethics Board review because it relies exclusively on publicly available information that is legally accessible to the public and appropriately protected by law.

### Summary of Updates

Interaction terms between stroke and wave (linear and quadratic) conditions were included to the mixed effects models to allow variations in I/ADL trajectories between the two groups (stroke survivors and stroke-free adults).

